# Rehabilitation needs and mortality associated with the Covid-19 pandemic: a population-based study of all hospitalised and home-healthcare individuals in a Swedish healthcare region

**DOI:** 10.1101/2021.04.30.21256372

**Authors:** Anestis Divanoglou, Kersti Samuelsson, Emer. Rune Sjödahl, Christer Andersson, Richard Levi

## Abstract

**Background:** This first report of the Linköping Covid-19 Study (LinCoS) aimed at determination of Covid-19-associated mortality, impairments, activity and participation limitations denoting rehabilitation needs four months after discharge from hospital.

**Methods:** An ambidirectional population-based cohort study including all confirmed Covid-19 cases admitted to hospital during 1/03-31/05 and those living in home healthcare settings identified through a regional registry and evaluated through medical records, including WHO Clinical Progression Scale (CPS). All patients discharged from hospital were followed-up by structured telephone interview at 4 months post-discharge. Respondents indicated any new or aggravated persisting problems in any of 25 body functions and 12 activity/participation items and rated them for impact on daily life.

**Findings:** Out of 734 hospitalised patients, 149 were excluded, 125 died, and 460 were alive at 4-month follow-up of whom 433 (94.1%) were interviewed. In total, 40% reported impairments and activity/participation limitations affecting daily life and warranted further multi-professional rehabilitation assessment, predominantly those with severe disease and a considerable proportion of those with moderate disease. Cognitive and affective impairments were equally common in all groups and were reported by 20-40% of cases. Limb weakness was reported by 31%, with CPS 7-9 being four times more likely to report this problem as compared to CPS 4-5. 26% of those working or studying reported difficulties returning to these activities, this being 3.5 times more likely in CPS 7-9 as compared to CPS 4-5. 25% reported problems walking >1 km, with CPS 7-9 over three times more likely to report this as compared to the other two sub-groups. 90-day mortality rate of Covid-19 associated deaths was 15.1%.

**Interpretation:** Most rehabilitation needs after Covid-19 involved higher cerebral dysfunction both in patients with moderate and severe disease. This should be considered when designing services aiming at minimizing long-term disability.

**Funding:** ALF grant and Region Östergötland.

## Background

This report presents population-based data from the first wave of Covid-19 in one of 21 healthcare regions in Sweden, adding much needed rehabilitation-focused evidence to existing studies. In contrast to other European and Nordic countries, Sweden followed a different approach towards adapting measures limiting the spread of the disease characterized by voluntary public health recommendations, as opposed to stringent lockdown measures.^1,2^ Region Östergötland (RÖ) is one of three regions where the pandemic hit particularly hard and early after the outbreak in the beginning of 2020. As of February 2021, the pandemic has shown a distinct biphasic course in this region, with the first phase occurring in the beginning of March, with a peak in April and May. After a gradual tailing off, a resurgence of a second wave occurred in November 2020.

Already in the first half of 2020, several reports on Covid-19 showed a wide array of neurological problems potentially associated to this infection.^3-5^ In relation to the topic of the present study, it has been reported that between 60-85% experienced at least one persisting symptom such as physical decline, fatigue, cognitive dysfunction, pain and sleep disturbance.^4,6-9^ Based on these findings, several authors have highlighted the importance of rehabilitation both in the sub-acute and long-term perspective.^4,9-12^ In accordance with the World Health Organisation (WHO) International Classification of Functioning, Disability and Health (ICF), enumeration of symptoms and impairments on the body function level is not sufficient in order to assess the health status of a person. The latter requires an evaluation of the impact of symptoms on the levels of body functions, activity and participation, respectively, as well as impact of environmental factors.^13,14^ Up to now, few studies have assessed these domains, so evidence concerning long-term rehabilitation needs related to major life areas such as work or studies, social life and well-being is needed.^14^

Further, available studies have focused on correlations between patient and disease severity descriptors as it pertains to likelihood for developing various symptoms, but not on the impact of those in the person’s daily life. An assessment of the impact on daily life is necessary in order to evaluate rehabilitation needs. Further, to date, the focus of extant larger cohort studies has been on somatic rather than cognitive symptoms,^8^ despite several early reports indicating that symptoms attributable to disorders of higher brain functions are prevalent after Covid-19.^4,15,16^ Last and foremost, there is an urgent need for population-based studies exploring long-term outcomes,^17^ as all available studies are single-or multi-hospital case series not capturing a defined region, and thus presenting with varying degree of selection bias.

To present a complete picture of the outcomes of a disease, it is also relevant to evaluate the disease-associated mortality. This is particularly difficult in the case of Covid-19, where fatal outcome has been shown to be strongly correlated with high age and frailty.^2,18,19^ This may risk inflating the Covid-19-associated mortality by including deaths from unrelated reasons. Most available studies report overall mortality, rather than evaluating the Covid-19-associated mortality. Bone et al ^20^ addressed these problems by using registry data and a statistical model to estimate Covid-19 deaths. A case-by-case assessment of the relevance of the infection to a fatal outcome would be of value.

In summary, and in accordance with a recently updated rapid living Cochrane systematic review on rehabilitation and Covid-19, there is a need for population-based studies that explore long-term outcomes in terms of Covid-19-associated mortality and rehabilitation needs.^21^ In particular, such assessments need to encompass not only body functions, but also activity and participation levels. In fact, only by assessing disease consequences in terms of all levels of functioning, as well as disease-associated mortality, can the outcomes of this pandemic be determined in a holistic manner.

With this in view, the main objective of this first report from the Linköping Covid-19 Study (LinCoS) was to determine Covid-19-associated mortality, as well as Covid-19–associated rehabilitation needs four months after discharge from hospital. The overall purpose was to inform the provision of care and rehabilitation in order to address the long-term sequels of the pandemic.

## Methods

### Study design

LinCoS comprises a planned series of sub-studies all stemming from a population-based cohort of patients hospitalised for Covid-19. LinCoS employs an ambidirectional design, by retrospective case identification and a prospective follow-up as regards long-term rehabilitation needs and mortality. The reporting is informed by the Strengthening the Reporting of Observational studies in Epidemiology (STROBE) statement for cohort studies.^22^ The primary research questions were:

- What are the frequencies, types and degrees of Covid-19-associated impairments and activity/ participation limitations at 4 months post-discharge?
- What is the Covid-19-associated mortality?

### Setting and inclusion criteria

As of December 2019, RÖ comprised 465.495 inhabitants.^23^ The region encompasses four hospitals, one of which is a university hospital (total capacity of 1,000 beds; 600 of which at the university hospital), as well as facilities designated to home healthcare. Regional home healthcare is administered in 136 institutions (care homes, with approximate capacity of 4,500 places) or, in a limited number of cases, in a patient’s home (approximately 2,500 individuals, with a proportion of them needing home care). In Sweden, the concept of home healthcare denotes consistent care of people unable to sustain independent living,^24^ predominantly comprising elderly, frail and/or individuals with dementia, as well as those with severe and chronic multi-morbidity with similar limitations. As a rule, these individuals are treated in a home healthcare setting rather than being hospitalised.

LinCoS included all registered inhabitants in RÖ with a laboratory confirmed Covid-19 diagnosis, and who also met either of the following criteria:

1. admission to hospital during the period March 1^st^ – May 31^st^ 2020 (“hospitalized patients”); or

2. living in a home healthcare setting during this period (“home healthcare individuals”).

The following exclusion criteria were implemented:

- for hospitalised patients – coincidental Covid-19 diagnosis as primary reason for hospitalisation (e.g. planned labour, elective or acute operation, other acute medical condition) and where Covid-19 did not extend or otherwise influence the clinical course;
- for home healthcare individuals – surviving >90 days after positive Covid-19 test;
- younger age than 15 years.

Additional exclusion criteria pertaining to research question related to rehabilitation needs were:

- for hospitalised patients – severe pre-existing comorbidities (e.g. severe dementia, terminal/ palliative care); and
- for home healthcare individuals – premorbid condition precluding hospitalisation for Covid-19 (e.g. severe dementia, terminal/ palliative care).

### Procedures

To identify hospitalised patients with Covid-19, all regional medical records during the study period were scrutinised. Two senior physicians (CA and RS) performed an audit of the medical records of all hospitalised patients with a laboratory confirmed Covid-19 diagnosis, and extracted key indicators pertaining to socio-demographic and clinical variables, including pre-morbid level of function. For quality assurance purposes, the other members of the inter-professional research team then verified this audit against corresponding medical records. The Panel presents the operational definitions used in this study.

To identify Covid-19 deaths in home healthcare, a regional population healthcare registry reporting to the national authorities was used for initial case identification. Medical record data pertaining to individuals thus identified were then scrutinized (CA, RS) to establish a pre-morbid level of function and to assess whether cause of death was associated with Covid-19.

All previously hospitalised patients alive at 4 months post-discharge were then contacted by an experienced rehabilitation professional (physician, neuropsychologist, occupational therapist, or physiotherapist) for a structured telephone interview to assess persisting Covid-19-associated symptoms and limitations in activity and participation. Interviews were conducted at a median (IQR) of 115 days (91-135) post-discharge. The interviews lasted 20-60 minutes, the longer duration required for interpreter-mediated interviews. For non-Swedish speaking respondents, in addition to the interviewer, a professional interpreter with expertise in the relevant language was also involved.

Interviews followed a standardised protocol comprising 37 questions, 25 on body function and 12 on activity/participation (Appendix 1). Questions reflected findings from research published in early 2020 regarding Covid-19-associated symptomatology,^25,26^ as well as previous experience of persisting problems among survivors of other viral diseases affecting the nervous system.^27^ Respondents were instructed to only report new or exacerbated problems in relation to Covid-19. For each such problem, the patient was asked to estimate its impact on everyday life on a scale from 1-5 (1: no impact; 2: to a minor degree; 3: to some degree; 4: to a high degree; 5: to a very high degree).

Self-rated health was assessed by asking respondents to rate their health status on a five-point Likert scale from very good to very bad.^28^ Respondents were asked to rate their health at the time of the interview and at the period immediately prior to infection as recalled. For assessment of persisting dyspnea, the Swedish version of the modified Medical Research Council (mMRC) Dyspnea Scale was used.^29^ This is a five-level scale relating self-rated dyspnea to physical activities, where level 0 corresponds to no or minimal problems even during strenuous exercise, and level 4 indicates severe breathlessness even in activities as modest as dressing or precluding leaving the home environment.^29^

Each interview was documented and evaluated by the interviewer in terms of rehabilitation needs. This evaluation was informed by number and types of reported problems, and their impact on daily life. Each evaluation was then discussed at regular multi-professional team conferences. Based on joint assessments from patient, interviewer and team on the current degree of Covid-19-associated disability, an appointment for subsequent individualised clinical assessment was made where appropriate.

After completion of data collection, we used the World Health Organisation (WHO) Clinical Progression Scale (CPS) and the corresponding data from the medical record, to classify the severity of Covid-19 in all included cases (Panel). ^30^ This version of the scale ranges from 0 (uninfected, no viral RNA detected) to 10 (death). It is worth noting that in the Swedish Covid-19 healthcare context, all mechanically ventilated cases are treated in an intensive care unit (ICU), but that ICU also admits patients in a critical condition who ultimately do not require mechanical ventilation.

### Statistical analysis

The assumption of normality was tested by visually inspecting histograms and by using the Shapiro-Wilk test. Descriptive data were presented as n (%), or as median and interquartile range (IQR) as data were skewed. No imputations were performed for missing data. Differences in proportions were examined by Chi^2^ test followed by post-hoc analysis of adjusted residuals. The Fisher Freeman Halton exact test was used when more than 20% of cells in a contingency table had an expected count less than 5. The Kruskal-Wallis test was used to compare differences between groups in continuous data, and in those cases where Kruskal-Wallis test was significant, the Bonferroni-Dunn’s test was used to adjust for multiple pairwise comparisons. Odds ratios and 95% confidence intervals were computed for between group differences. Statistical significance was set at alpha <0.05. Statistical analyses were performed with IBM SPSS statistics for Windows (Version 27, Chicago, Illinois, USA).

### Ethics committee approval

The Swedish Ethical Review Authority approved the study protocol (Dnr 2020-03029 and 2020-04443).

### Role of funding source

The study was funded by the ALF grant and Region Östergötland. The funder of the study had no role in study design, data collection, data analysis, data interpretation, or writing of the report. The corresponding authors (AD and RL) had full access to the full data in the study and accept responsibility to submit for publication.

**Panel. Operational definitions**

##### Total LOS

length of hospitalisation (in days) pertaining to Covid-19-related care. Total LOS was further divided into days in ICU and days in other wards.

##### Pre-morbid level of function

individual patient performance status and frailty during a month preceding Covid-19, as reflected in the medical record in terms of employment, mobility and impact (type, severity and control) of co-morbidities. This assessment was summarised in a modification of the WHO/ECOG Performance Status^31^ and the Frailty Score according to Rockwood^32^ to yield an assignment of the pre-morbid level of function in either of four groups:

1. No or mild frailty: no restriction in daily life activities.
2. Moderate frailty: person mobile and autonomous, but unable to perform physically demanding activities/ work.
3. Considerable frailty: person usually able to perform basic daily life activities, but periodically confined to bed or chair.
4. Severe frailty: person permanently unable to perform activities of daily living and/or confined to bed or chair, e.g. dementia necessitating care.

##### Hospitalised death

fatalities of any hospitalised patients meeting inclusion criteria, dying either during hospital care or after discharge.

##### Home healthcare death

fatalities occurring among individuals with Covid-19 in home healthcare without prior admittance to hospital for Covid-19-related care. This category did not include individuals in home healthcare who were admitted to hospital and subsequently died within the study period. Such individuals were assigned to the hospitalised deaths category.

##### Covid-19-associated death

fatalities where Covid-19 was assessed as a dominating or contributing factor to outcome. That assessment was performed by considering death certificate diagnoses in relation to preceding events as documented in the medical record. The 30- and 90-day mortality after positive Covid-19 test were assessed for both hospitalised and home healthcare individuals.

##### Severity of Covid-19

the highest WHO Clinical Progression Scale (CPS) grade during hospitalisation and/ or follow-up time. In subsequent analysis, and in accordance with this scale, a highest score of WHO CPS 4 and 5 was denoted as “hospitalised moderate disease”, a highest score of grade 7-9 was denoted as “hospitalized severe disease”, and a highest score of grade 10 denoted “fatal outcome”.^30^ In addition, we chose to report WHO CPS 6 (“hospitalised, severe disease, non-mechanically ventilated”) as a separate sub-group.

##### Problem affecting daily life

the subset of all persisting problems reported at the interview that were evaluated by the patient as having “some, high or very high impact” on their daily life. For the purposes of assessing rehabilitation needs, problems reported as having “no, or minor impact” on daily life, respectively, were excluded from subsequent analysis in this study.

## Findings

### Cohort

In total, 734 patients were hospitalised with Covid-19 during the study period. Out of these, 77 were excluded from this study, as their hospitalisation was coincidental to Covid-19. Overall, 657 patients were thus hospitalised due to Covid-19. In addition, 110 individuals in home healthcare settings, not admitted to hospital, were diagnosed with Covid-19 and died within 90 days after positive test. Figure 1 presents a flowchart of the study population.

**Figure.**
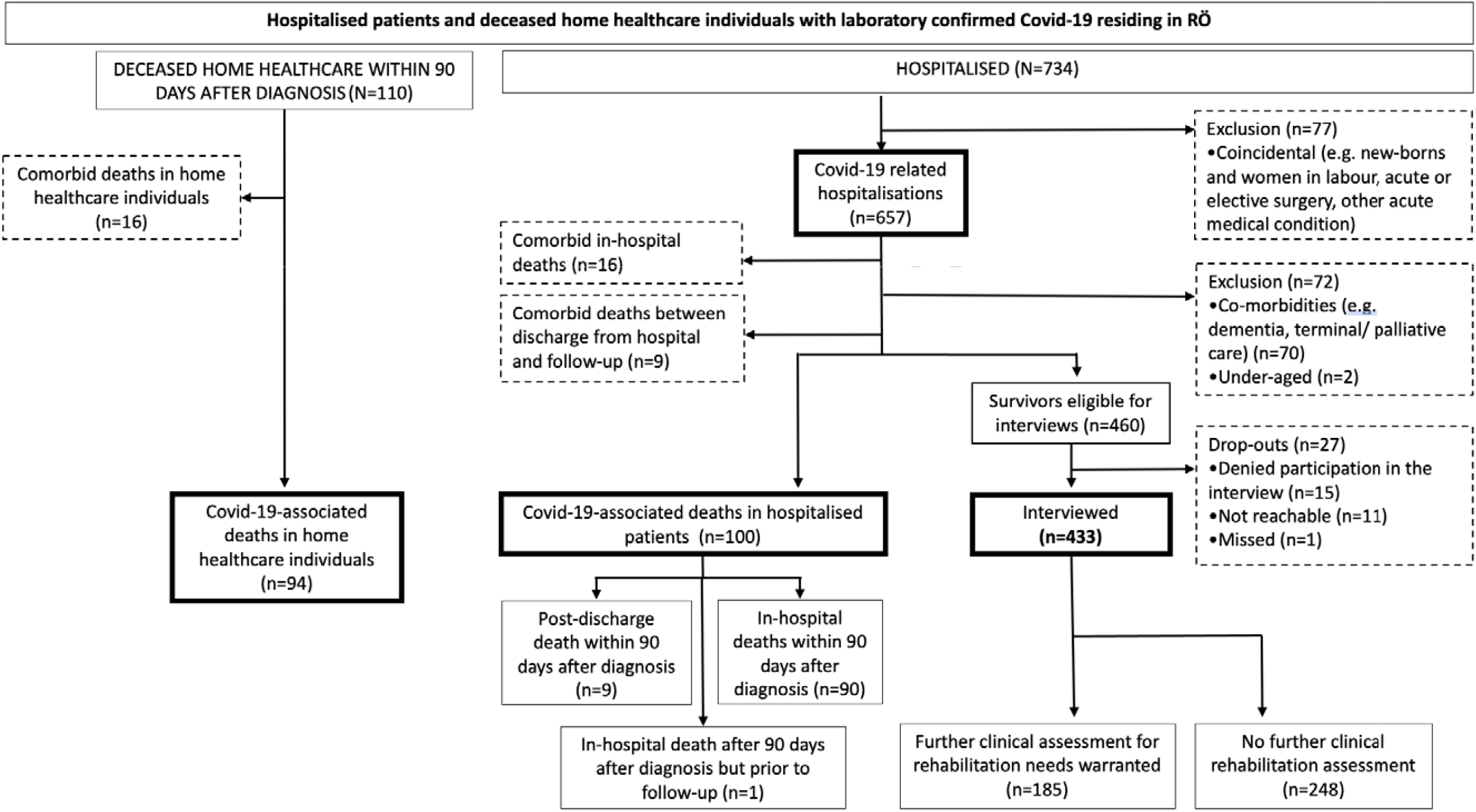

Flowchart of hospitalised (admission during study period) patients and deceased home healthcare individuals (positive test dur ing study period) with laboratory confirmed Covid-19 in Region Östergötland. Study period: March 1^st^ to May 31^st^ 2020.

### Survivors

Out of 460 previously hospitalised patients alive at 4-month follow-up, 433 were interviewed (response rate 94·1%). The majority of respondents (78·3%) had had moderate disease as assessed with WHO CPS; 133 (30·7%) had been grade 4 and 206 (47·6%) grade 5. Of those with severe disease, 38 cases (8·8%) had been grade 6, no case grade 7, 24 cases (5·5%) grade 8 and 32 cases (7·4%) grade 9. Hospital LOS was longer for those with grade 7-9 as compared to grade 4-5 (p<·001) and grade 6 (p=0·014).

### Impairments in body functions and activity/ participation limitations at follow-up

13% did not report any new or aggravated problems. 44·3% reported problems but not to an extent requiring multi-professional rehabilitation assessment. The remaining 42·7% reported problems affecting daily life to a concerning degree and were invited for clinical rehabilitation assessment.

Frequencies and ORs for impairments in body functions and activity/participation limitations affecting daily life are presented in Table 3. The most common body function impairments were mental fatigue (38%) and limb weakness/fatiguability (31%). As for activity and participation limitations, the most common problems were difficulty being physically active (35%) and difficulty managing work or studies (26%).

**Table 1.** Demographic characteristics and premorbid level of function of enrolled patients. Premorbid level of unction (1-4): (1) No or mild frailty: no restriction in daily life activities; (2) Moderate frailty: person mobile and autonomous, but unable to perform physically demanding activities/ work; (3) Considerable frailty: person usually able to perform basic daily life activities, but periodically confined to bed or chair; (4) Severe frailty: person permanently unable to perform activities of daily living and/or confined to bed or chair, e.g. dementia necessitating care.

|  | Hospitalised<br>Total group<br>(N=433) | Hospitalised Interviewed Survivors (n=433) |  |  | Hospitalised 90-day Covid-19<br>associated death | Home healthcare<br>90-day Covid-19<br>associated death |
| --- | --- | --- | --- | --- | --- | --- |
|  |  | WHO CPS 4-5<br>(n=339) | WHO CPS 6<br>(n=38) | WHO CPS 7-9<br>(n=56) | WHO CPS 10<br>(n=99) | WHO CPS 10<br>(n=94) |
| Age in years –<br>median (IQR) | 61.0 (49.0-74.0) | 61.0 (49.0-75.0) | 60.0 (52.8-72.3) | 62.0 (56.3-69.0) | 81.0 (73.0-88.0)* | 88.0 (84.0-93.0)* |
| Sex |  |  |  |  |  |  |
| Males | 246 (56.8%) | 176 (51.9%)* | 27 (71.1%) | 43 (76.8%)* | 52 (52.5%) | 40 (42.6%) |
| Females | 187 (43.2%) | 163 (48.1%) | 11 (28.9%) | 13 (23.2%) | 47 (47.5%) | 54 (57.4%) |
| Premorbid level of<br>function (1-4) |  |  |  |  |  |  |
| 1 | 172 (39.7%) | 132 (38.9%) | 16 (42.1%) | 24 (42.9%) | 1 (1.0%) | 0 |
| 2 | 135 (31.2%) | 97 (28.6%) | 13 (34.2%) | 25 (44.6%) | 17 (17.2%)* | 0* |
| 3 | 117 (27.0%) | 102 (30.1%) | 8 (21.1%) | 7 (12.5%) | 41 (41.4%)* | 3 (3.2%)* |
| 4 | 8 (1.8%) | 7 (2.1%) | 1 (2.6%) | 0 | 38 (38.4%)* | 91 (96.8%)* |
| Missing | 1 (0.2%) | 1 (0.3%) | 0 | 0 | 2 (2.0%) | 0 |
| Data presented as n (%) unless otherwise specified<br>* p≤0.05; comparisons performed between the three WHO CPS groups, and also between hospitalised and home healthcare deaths. |  |  |  |  |  |  |

**Table 2.** Clinical process indicators and summarised interview outcomes for hospitalised patients.

|  | Hospitalised<br>Interviewed Survivors<br>Total group<br>(n=433) | Hospitalised Interviewed Survivors |  |  | Hospitalised 90-day<br>Covid-19 associated death |
| --- | --- | --- | --- | --- | --- |
|  |  | WHO CPS 4-5<br>(n=339) | WHO CPS 6<br>(n=38) | WHO CPS 7-9<br>(n=56) | WHO CPS 10<br>(n=99) |
| LOS Total, in days | 6.0 (3.0-13.0) | 4.0 (2.0-8.0)* | 13.0 (10.0-18.5)* | 36.0 (23.0-55.0)* | 7.0 (4.0-11.0) |
| ICU admission, n (%) | 68 (15.7%) | 4 (1.2%)* | 8 (21.1%) | 56 (100.0%)* | 15 (15.2%) |
| LOS ICU, in days | 15.0 (8.0-25.8) | 1.0 (1.0-1.0) | 1.0 (1.0-2.8) | 17.5 (10.3-29.0)** | 16 (5.0-23.0) |
| Duration of mechanical<br>ventilation, in days | 15.5 (10.0-24.5) | 0 | 0 | 15.5 (10.0-24.5)** | 16.0 (6.0-23.0) |
| Time between discharge<br>and follow-up, in days | 115.0 (91.0-135.5) | 116.0 (91.0-143.0) | 105.0 (94.5-122.5) | 114.0 (89.0-133.8) | N/A |
| Number of problems<br>affecting daily life, n (%) |  |  |  |  | N/A |
| 0 | 132 (30.5%) | 111 (32.7%) | 13 (34.2%) | 8 (14.3%) |  |
| 1-5 | 163 (37.6%) | 129 (38.1%) | 13 (34.2%) | 21 (37.5%) |  |
| 6-10 | 63 (14.5%) | 47 (13.9%) | 2 (5.3%) | 14 (25.0%) |  |
| 11-20 | 51 (11.8%) | 35 (10.3%) | 6 (15.8%) | 10 (17.9%) |  |
| >20 | 24 (5.5%) | 17 (5.0%) | 4 (10.5%) | 3 (5.4%) |  |
| Further clinical<br>assessment for<br>rehabilitation needs, n (%) | 185 (42.7%) | 122 (36.0%)* | 18 (47.4%) | 45 (80.3%)* | N/A |
| Data are presented as median (IQR) other otherwise specified; ICU: Intensive care unit; LOS: Length of stay; WHO CPS: World Health organisation Clinical progression scale; N/A: not applicable; * indicates p≤0.05; comparisons performed between the three WHO CPS groups, and also between hospitalised and home healthcare deaths. |  |  |  |  |  |

**Table 3.**
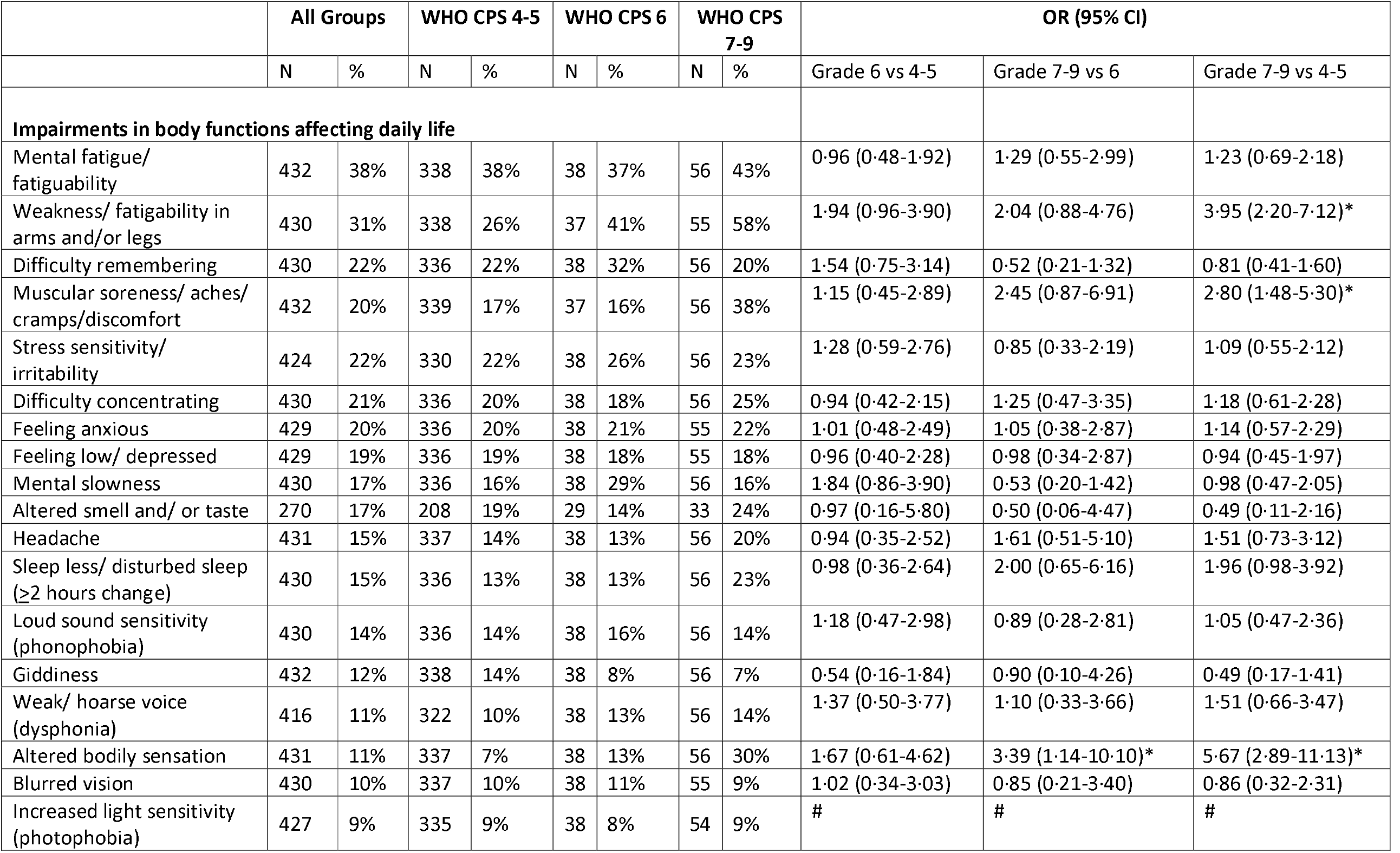
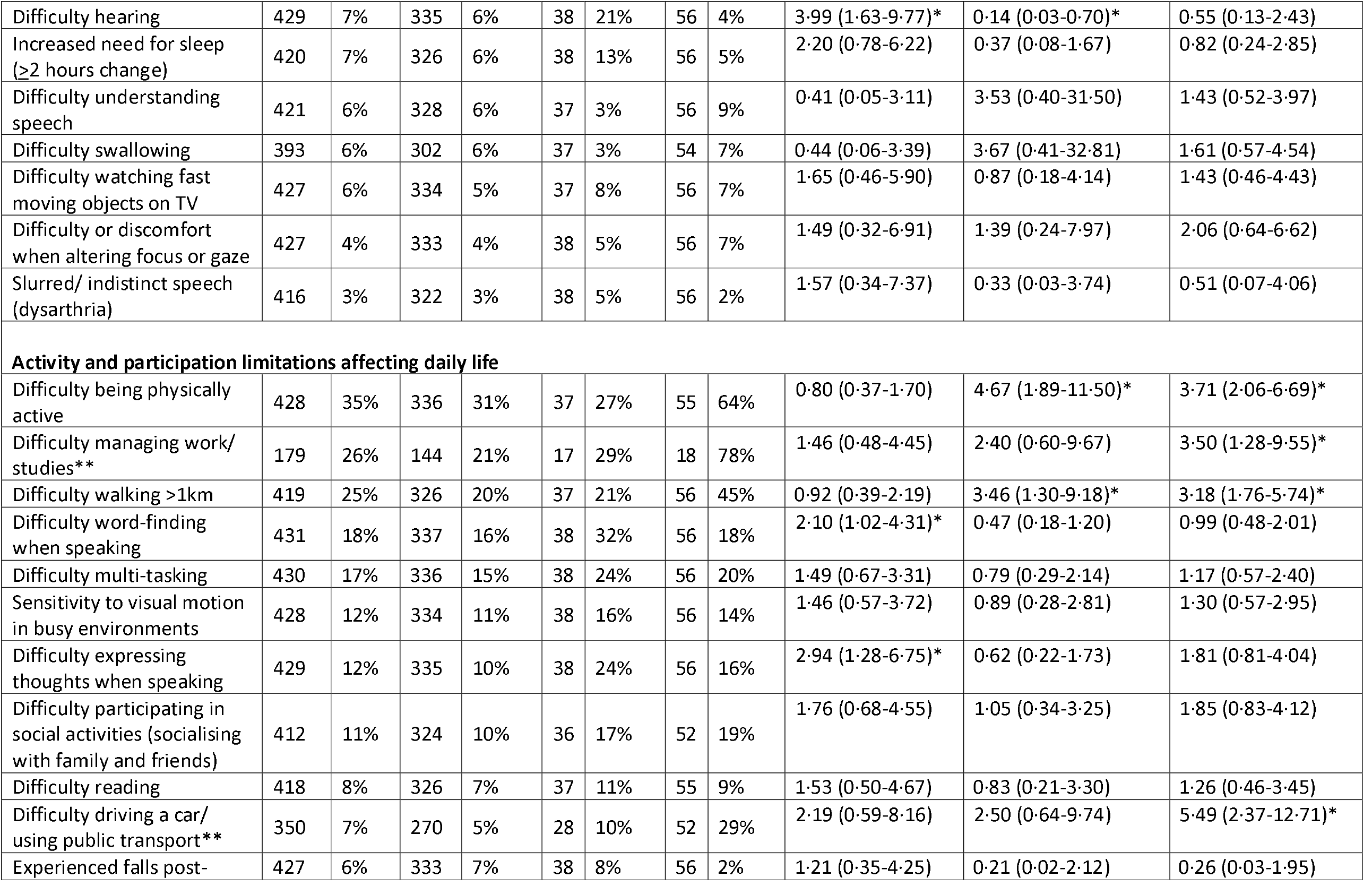
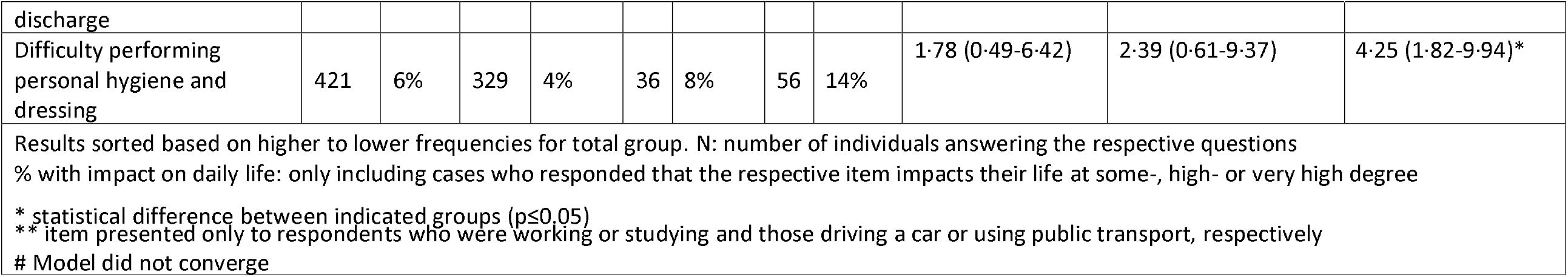
Distribution of reported symptoms at 4-month follow-up organised according to the WHO ICF impairments (body functions), and activity and participation limitations affecting daily life.

### Common problems affecting daily life across all WHO CPS sub-groups

On the body function (impairment) level, cognitive impairments were equally common across all groups. In total and in each sub-group, 40% reported mental fatigue, and at least 20% reported difficulties concentrating and remembering. Affective impairments such as feeling anxious, stressed or depressed were likewise reported by over 20% across all groups. Limb weakness was reported by 31% of the whole group, but with grade 7-9 being four times more likely to report this problem as compared to the grade 4-5 sub-group (OR: 3·95, 95% CI: 2·20-7·12).

On the activity and participation level, nearly 20% of the total group reported difficulties multi-tasking. A quarter of those working or studying prior to Covid-19 hospitalisation reported difficulties returning to these activities, with grade 7-9 being 3·5 times more likely to report such problems as compared to grade 4-5 (OR: 3·50, 95% CI: 1·28-9·55). More than one third of the total group reported not being able to be as physically active as previously, with grade 7-9 approximately four times more likely to report such problems as compared to grade 4-5 (OR: 3·71, 95% CI: 2·06-6·69) and grade 6 (OR: 4·67, 95% CI: 1·89-11·50) respectively. Furthermore, one quarter of the total group reported problems walking more than 1 km, again with grade 7-9 being over three times more likely to report this problem as compared to the other two groups (OR: 3·18, 95% CI: 1·76-5·74; OR: 3·46, 95% CI: 1·30-9·18).

### Common problems affecting daily life in specific WHO CPS sub-groups

#### On the body function (impairment) level

grade 7-9 was almost 4 times more likely to report muscular cramps/soreness/aches/discomfort than grade 4-5 (OR: 2·80, 95% CI: 1·48-5·30). Furthermore, grade 7-9 was almost six and three times respectively more likely to report altered body sensation than grade 4-5 (OR: 5·67, 95% CI: 2·89-11·13) and grade 6 (OR: 3·39, 95% CI: 1·14-10·10). Grade 6 was four and seven times more likely to report hearing problems as compared to grade 4-5 (OR: 3·99, 95% CI: 1·63-9·77) and grade 7-9 (OR: 7·20, 95% CI: 1·44-36.11) respectively. Grade 6 was also three times more likely to report difficulties with expressing thoughts when speaking as compared to grade 4-5 (OR: 2·94, 95% CI: 1·28-6·75).

#### On the activity and participation level

grade 7-9 was 5.5 times more likely to report difficulties driving a car or using public transport as compared to grade 4-5 (OR: 5·49; 95% CI: 2·37-12·71).

### Less common, but concerning problems affecting daily life across WHO sub-groups

On the activity and participation level, 6% of the total group and 14% of grade 7-9 reported difficulties undertaking personal hygiene and dressing activities. Difficulties participating in social activities were reported by 11% of the total group, and as high as 17% and 19% in grade 6 and 7-9, respectively.

### Perceived breathlessness

Overall, 24·3% of the 420 respondents reported no breathlessness even at strenuous exercise at follow-up. Conversely, approximately 20% of the total group reported scores suggesting a considerable activity limitation (scores 3 and 4). The frequencies of individual responses are shown in table 4. There were no in-between group differences in regard to perceived breathlessness.

**Table 4.** Modified Medical Research Council (mMRC) Dyspnea Scale at 4-month follow-up, and Self-rated health difference between prior to admission and 4-month follow-up.

|  | All groups<br>N=420 | WHO CPS 4-5<br>N=327 | WHO CPS 6<br>N=37 | WHO CPS 7-9<br>N=56 |
| --- | --- | --- | --- | --- |
| **mMRC score at 4-month follow-up** |  |  |  |  |
| **0** | 102 (24.3%) | 85 (26.0%) | 10 (27.0%) | 7 (12.5%) |
| **1** | 148 (35.2%) | 110 (33.6%) | 11 (29.7%) | 27 (48.2%) |
| **2** | 82 (19.5%) | 67 (20.5%) | 9 (24.3%) | 6 (10.7%) |
| **3** | 51 (12.1%) | 36 (11.0%) | 5 (13.5%) | 10 (17.9%) |
| **4** | 37 (8.8%) | 29 (8.9%) | 2 (5.4%) | 6 (10.7%) |
| **Self-rated health difference between the period prior to admission and 4-month follow-up** |  |  |  |  |
|  | All groups<br>N=427 | WHO CPS 4-5<br>N=334 | WHO CPS 6<br>N=38 | WHO CPS 7-9<br>N=55 |
| **Not worsened** | 238 (55.7%) | 197 (59.0%) | 22 (57.9%) | 19 (34.5%) |
| **Worsened** | 189 (44.3%) | 137 (41.0%)* | 16 (42.1%) | 36 (65.5%)* |
| * statistical difference between indicated groups ($p \leq 0.05$); WHO CPS: World Health Organisation Clinical Progression Scale | | | | |

### Self-rated health

About half of all respondents (56%) at follow-up reported no worsening in self-rated health at 4 months after discharge as compared with their recalled health status before Covid-19 (Table 4). However, 44% reported a poorer state of health. This was more common in grade 7-9 as compared to grade 4-5 (p=0.003).

### Mortality

Out of the 657 patients hospitalised due to Covid-19, 90 died in hospital from reasons associated with Covid-19, yielding an in-hospital Covid-19-associated mortality rate of 13.7% (95% CI: 10·9-16·5). The 30-and 90-day Covid-19-associated mortality rate was 14·6% (95% CI: 11·7-17·5) and 15·1% (95% CI: 11·3-18·0) respectively, also including those dying after discharge from hospital.

In regard to home healthcare individuals with a Covid-19 associated death, 93 died within 30 days after diagnosis (94 individuals within 90 days after diagnosis). When comparing the latter group with the corresponding 90-day mortality group of hospitalised patients, hospitalised patients were younger (81 years versus 88 years), and with superior pre-morbid levels of function (Table 1). Both deceased home healthcare individuals and hospitalised patients died after a median (IQR) of 7 (5-13) and 8 days (5-13) after Covid-19 diagnosis, respectively.

## Discussion

This study provides population-based outcomes associated with Covid-19 at 4 months follow-up in cases either being hospitalized due to this disease or being cared for in home healthcare institutions. It excluded cases with coincidental Covid-19 diagnosis and cases with severe pre-existing comorbidities where rehabilitation was precluded. Thus, a more correct assessment of actual rehabilitation needs was yielded.

The current study found that 44% of patients previously hospitalised for Covid-19 reported worsened health status at 4-month follow-up as compared to the period prior to Covid-19, with those with severe disease (as defined by WHO CPS grade 7-9) more likely to report such worsening as compared to those with moderate disease severity. At follow-up, approximately 20% reported dyspnea scores indicating considerable activity limitations, independently of disease severity. Cognitive and affective impairments were reported by 20-40% of cases and were equally common in all disease severity sub-groups. 31% reported limb weakness, 26% of those working or studying reported difficulties returning to these activities, and 25% reported problems walking >1 km, with those with grade 7-9 more likely to report these problems as compared to those with moderate disease. Among hospitalised patients, the 90-day (after diagnosis) mortality rate associated with Covid-19 was 15.1%.

There are currently few studies pertaining to long-term outcomes of the Covid-19 pandemic. Our study is fundamentally in agreement with existing studies regarding symptom enumeration.^4,6,33^ The main contribution of this study lies in its focus not primarily on symptom enumeration, but on identifying the Covid-19 associated rehabilitation needs resulting from diverse impairments and activity/participation limitations. Problems at all levels of functioning comprise the impetus to rehabilitation. Few studies provide this scope.^4,11^ The use of the WHO ICF taxonomy, not utilized in studies on outcomes of Covid-19 hitherto, characterises our approach. Additionally, mortality is one key outcome not to be excluded from any analysis of consequences of a disease. This provides a backdrop in relation to which outcomes are put in context.

The most relevant study to date in this context evaluated a cohort of 1,733 individuals in Wuhan, China, at 6 months post-discharge.^8^ This single-hospital study reported a similar spectrum of impairments, corroborating our findings of symptoms attributable to several organ systems. However, that study did not include assessment of neuropsychological dysfunctions, neither impact of any symptom on daily life. As compared to the present study, it reported considerably lower prevalence of persisting problems related to mobility, personal care and performance of usual activities as assessed solely by EQ-5D.^8^ This may have been influenced by very few patients receiving mechanical ventilation (only 10 out of 1,733; 0·5%) as compared to 13% in this study and other similar large studies. ^2,26^

In addition to giving frequencies of problems, this study addresses rehabilitation needs. Indicators of complex rehabilitation needs include: a multitude of problems affecting daily life; problems affecting several functional domains; and/or problems considered red flags from a rehabilitative perspective, e.g. ambulation difficulties, dysphagia and inability to perform personal care.

It has been reported that patients needing mechanical ventilation experience more severe impairments and activity/participation limitations as compared to moderately ill patients.^2^ Our results support this notion, especially in relation to difficulties being physically active, walking longer distances, driving and using public transportation and, perhaps foremost, difficulties returning back to work or studies. Persisting somato-sensory and somato-motor impairments, possibly related to critical illness myopathy (CIM) and/or critical illness polyneuropathy (CIP) were reported almost exclusively among the patients having required ICU treatment in the acute phase. In fact, 80% of patients in grade 7-9 were in need of further multi-professional clinical rehabilitation assessment.

Such a need for further assessment was also present in almost half of patients receiving NIV/Optiflow and in over one third of cases with only oxygen or no-oxygen required. Thus, it is necessary to evaluate rehabilitation needs for all hospitalised Covid-19 cases. There was a clear preponderance of neurocognitive and, more generally, neuropsychological problems. This again holds true not only for ICU-treated patients, but also for patients not requiring mechanical ventilation. The most common impairment at follow-up was mental fatigue which was reported to a similar extent regardless of disease severity.

Overall, 44% reported decline in self-rated health persisting at follow-up. Grade 7-9 more commonly reported such decline as compared to grade 4-5. The few studies reporting such a comparison show a similar pattern but to a lesser extent. ^4^

In-hospital mortality rate in the present study was 13·7%, considerably lower than corresponding figures in other large studies reporting mortality between 21-40%.^18,19,26^ In our study, 14·5% (16/110) of home healthcare deaths and 20% (25/125) of hospitalised deaths were evaluated as not being Covid-19 associated. A recent study used statistical modelling and reported 26·1% additional deaths, i.e. deaths not attributed to Covid-19. ^20^ We put effort in assessing the contributory role of Covid-19 on occurrence of death, and this may have contributed to our lower mortality rate. Controlling for non-associated deaths is obviously of importance, as fatal outcomes strongly correlate with high age, severe comorbidities and in particular premorbid frailty, factors which by themselves cause mortality. ^18,19^

On the other hand, if only hospital mortality is considered, this would lead to an underestimation of mortality associated with Covid-19, as the majority of home healthcare individuals dying with Covid-19 never were admitted to hospital. Thus, similar to other studies, ^20^ we chose to include individuals with a confirmed diagnosis dying both in hospital as well as in home healthcare settings. Thereby, we applied similar criteria in both groups for associating death to Covid-19. In conclusion, in line with other studies, ^18,26,34^ fatal outcomes occurred predominantly, but not exclusively, among patients with severe premorbid frailty, partially related to high age, partially to comorbidities. However, even when considering these factors, during the study period 99 hospitalised patients and an additional 94 home healthcare individuals died due to Covid-19 within 90 days after their positive test.

In regard to study limitations, due to the state of affairs in our country during the period under study, only hospitalised patients and individuals in home healthcare were consistently diagnosed by viral testing. Therefore, a probably large number of undiagnosed and non-hospitalised cases of Covid-19 could not be assessed in this study as regards persisting rehabilitation needs. It is very likely that, for this reason, this study provides a modest estimate for the true magnitude of rehabilitation needs.

Further, rehabilitation needs associated with Covid-19 were not assessed in individuals where their premorbid condition was so poor as to preclude rehabilitation initiatives. In particular, individuals in home healthcare suffer a very poor health condition due to old age, dementia and/or other major disabling disorders, as this is the main reason to live in such facilities. As a group they were not considered eligible for rehabilitation in the sense explored in this study, and were not assessed for that purpose. Last, the generalisability of our findings may be limited in countries with distinctly different healthcare systems, socioeconomic structure and culture, given that these aspects may influence outcomes after Covid-19.

## Conclusion

In contra-distinction to early conceptions that Covid-19 predominantly is a respiratory tract disorder also in terms of long-term consequences, most rehabilitation needs at four months after discharge involved higher cerebral dysfunction. This has to be considered when designing services aiming at minimizing long-term disability associated to Covid-19.

## Data Availability

As subsequent follow-up investigations related to LinCoS are in progress, data presented in this report will not be made available to others at this stage. After completion of LinCoS, data can be made available to others upon request and after an individualised assessment by the LinCoS Project Group.

## Authors’ contributions

RL with assistance by AD and KS conceptualised the idea. RL, AD and KS designed the study and received assistance from CA and RS. AD had the primary responsibility for data curation, and was assisted by KS, CA and RS. AD administered the project. AD performed most data analysis and was assisted by KS. RL, KS and AD applied for funding. AD and RL wrote the original draft, and together with KS, CA and RS edited subsequent versions of the draft until finalisation of the manuscript. RL and AD were involved in the investigation process including data collection.

## Acknowledgements

We acknowledge all collaborators at the Department of Rehabilitation Medicine in Linköping, and in particular the leadership team of the department, for facilitating the integration of research in the clinical setting. Last, we acknowledge Professor Michelle Chew, MD, PhD at the Department of Anaesthesiology and Intensive Care in Linköping for providing valuable assistance with interpretation of ICU data.

## Research in context

### Evidence before this study

We searched PubMed with no language restriction for studies exploring mortality and long-term consequences after hospitalisation related to Covid-19 up to Jan 15, 2021. We used search terms (“SARS-CoV-2” OR “Covid-19”) AND (“hospital*”) AND (“mortality” OR “consequences” OR “follow-up” OR “long-term”) and their synonyms. Studies reported a diverse array of symptoms from many areas of body function, and that frequencies of such symptoms were high. Additionally, mortality was widely reported to be considerably higher than in most previous viral pandemics, e.g. influenza.

However, results have not translated into actual outcomes in terms of rehabilitation needs. This is due to several factors, the most important of which are a mere enumeration of symptoms in the absence of assessment in terms of corresponding activity and participation limitations, and provision of outcomes in the absence of evaluation of the degree to which reported outcomes indeed could be attributed to Covid-19 rather than to pre-and comorbid conditions. More recent studies have made attempts to expand on functional consequences, but without implementation of the WHO ICF taxonomy, which was specifically developed for the purpose of assessment and description of rehabilitation needs.

### Added value of this study

The findings of this study are based on a total regional cohort comprising individuals both hospitalised and residing in home healthcare, with 30-and 90-day outcomes as regards mortality, and 4-month post-discharge outcomes in survivors previously hospitalised due to Covid-19. Efforts were made to isolate the actual contribution of Covid-19 on outcomes, by assessing premorbid health status, comorbidities, coincidental cases, and by specifically documenting only new or aggravated symptoms occurring during and after Covid-19. Furthermore, respondents were asked to grade the relative impact on daily life for each such symptom, which in conjunction with questions specifically pertaining to activity and participation in accordance with WHO ICF provided results focused on actual rehabilitation needs associated with Covid-19.

### Implications of all the available evidence

The implications of this study are: 1. 40% of previously hospitalised survivors present significant rehabilitation needs impacting their daily life even at 4-months post-charge; 2. Such rehabilitation needs are most common among those with severe disease, i.e. had received mechanical ventilation, but were also present in about one third of patients classified as moderate disease; 3. Impairments in body function, activity and participation were reported in several domains, but in particular related to the nervous system; 4. Mortality associated with Covid-19 was considerable even after controlling for premorbid severe health problems and coincidental causes of morbidity and death. Our study found the Covid-19 pandemic to be a major challenge to rehabilitation services in particular, in order to minimize long-term disability caused by this pandemic.

