## Appendices for "Rehabilitation needs and mortality associated with the Covid-19 pandemic: a population-based study of all hospitalised and home-healthcare individuals in a Swedish healthcare region"

**Appendix I**

**Interview questions**

**Section A**

Consider the following question for each of the below items.

“At this moment in time, do you experience that the issue below comprise a new or aggravated problem as compared to prior to Covid-19 infection?”

If yes, to what extent does this new or aggravated problem affect your daily life?

1: no impact; 2: to a minor degree; 3: to some degree; 4: to a high degree; 5: to a very high degree

- Weakness/fatigability in arms and/or legs
- Difficulty walking >1km
- Difficulty being physically active
- Experienced falls post-discharge
- Altered bodily sensation
- Muscular soreness/ aches/ cramps/ discomfort
- Difficulty swallowing
- Altered smell and/ or taste
- Difficulty hearing
- Blurred vision
- Difficulty watching fast moving objects on tv
- Difficulty or discomfort when altering focus or gaze
- Increased light sensitivity (photophobia)
- Loud sound sensitivity (phonophobia)
- Headache
- Giddiness
- Sleep less/ disturbed sleep (≥2 hours change)
- Increased need for sleep (≥2 hours change)
- Mental fatigue/ fatiguability
- Stress sensitivity/ irritability
- Feeling anxious
- Feeling low/ depressed
- Difficulty concentrating
- Difficulty multi-tasking
- Mental slowness
- Difficulty remembering
- Difficulty understanding speech
- Difficulty word-finding when speaking
- Difficulty expressing thoughts when speaking
- Slurred/ indistinct speech (dysarthria)
- Weak/ hoarse voice (dysphonia)
- Sensitivity to visual motion in busy environments
- Difficulty participating in social activities (socialising with family and friends)
- Difficulty managing work/ studies
- Difficulty driving a car/ using public transport*****
- Difficulty performing personal hygiene and dressing
- Difficulty reading

**Section B**

modified Medical Research Council (mMRC) Dyspnea Scale

0 - “I only get breathless with strenuous exercise”

1 - “I get short of breath when hurrying on the level or walking up a slight hill”

2 - “I walk slower than people of the same age on the level because of breathlessness or have to stop for breath when walking at my own pace on the level”

3 - “I stop for breath after walking about 100 yards or after a few minutes on the level”

4 - “I am too breathless to leave the house” or “I am breathless when dressing”

Section C

a. At the moment, how would you describe your health status?

Very good; Good; Moderate; Bad; Very bad

b. At the period immediately prior to Covid-19 infection, how would you describe your health status?

Very good; Good; Moderate; Bad; Very bad

**Appendix II**

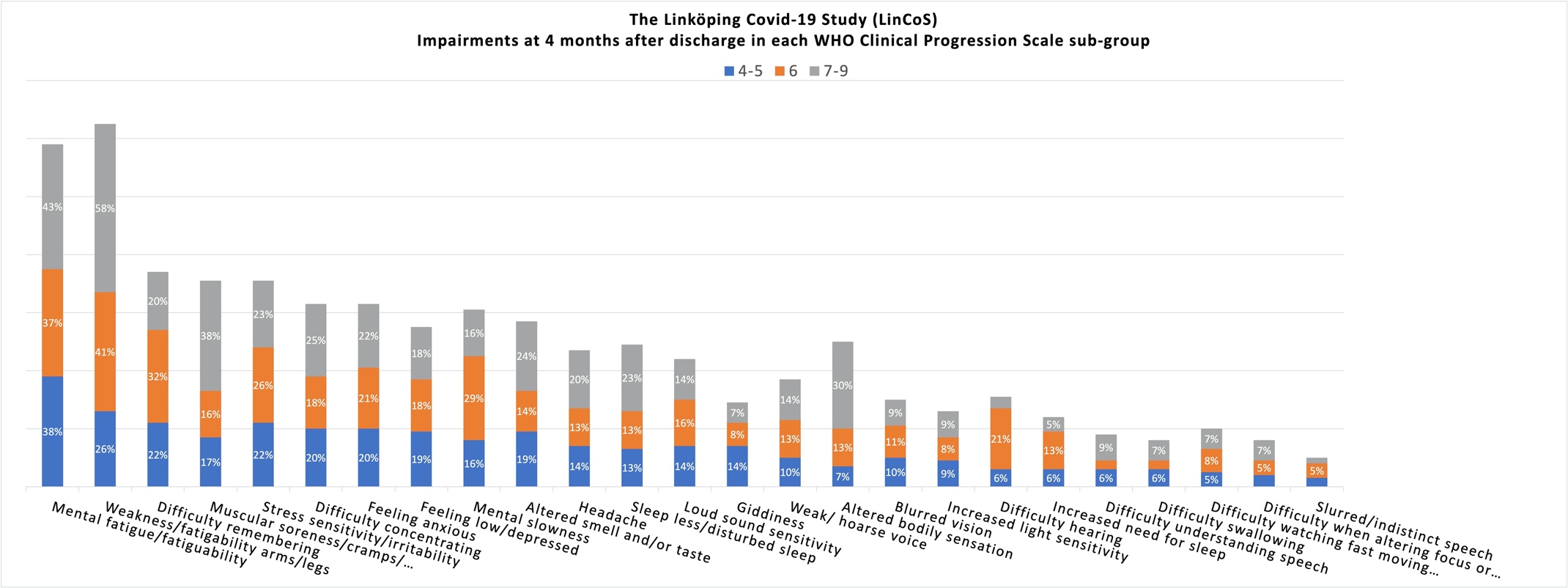

**Appendix II-Table 1.** Impairments affecting their daily life at 4 months after discharge - percentages of patients in each World Health Organization (WHO) Clinical Progression Scale (CPS) sub-group reporting. WHO CPS: grades 4-5 (hospitalised moderate disease, no oxygen or oxygen by mask or nasal prongs); grade 6 (hospitalised, severe disease, oxygen by NIV or high flow); grades 7-9 (hospitalised, severe disease, mechanical ventilation).

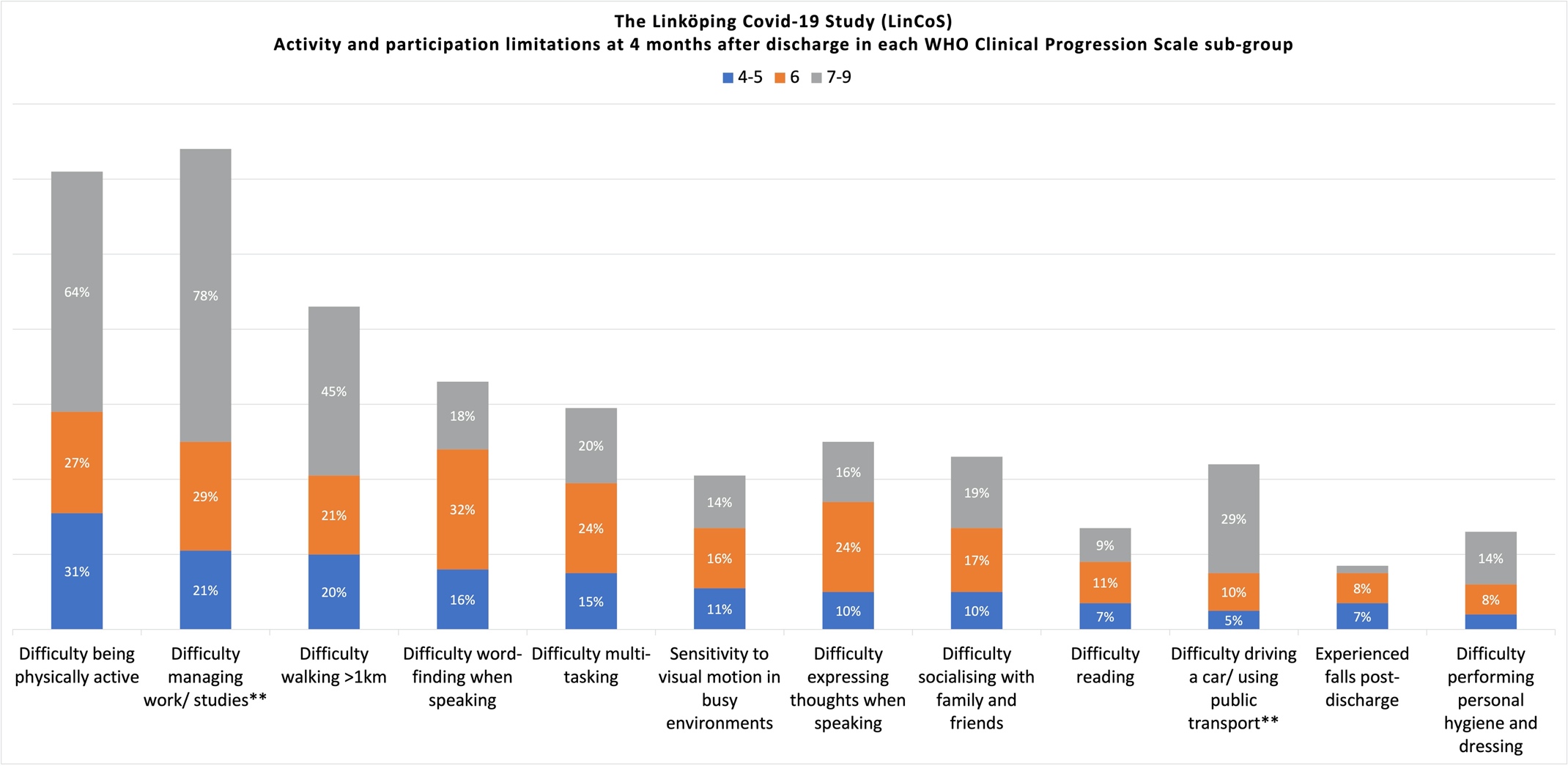

**Appendix II-Table 2.** Activity/participation limitations affecting their daily life at 4 months after discharge - percentages of patients in each World Health Organization (WHO) Clinical Progression Scale (CPS) sub-group reporting. **Answered only by patients for whom the question was relevant. WHO CPS: grades 4-5 (hospitalised moderate disease, no oxygen or oxygen by mask or nasal prongs); grade 6 (hospitalised, severe disease, oxygen by NIV or high flow); grades 7-9 (hospitalised, severe disease, mechanical ventilation).

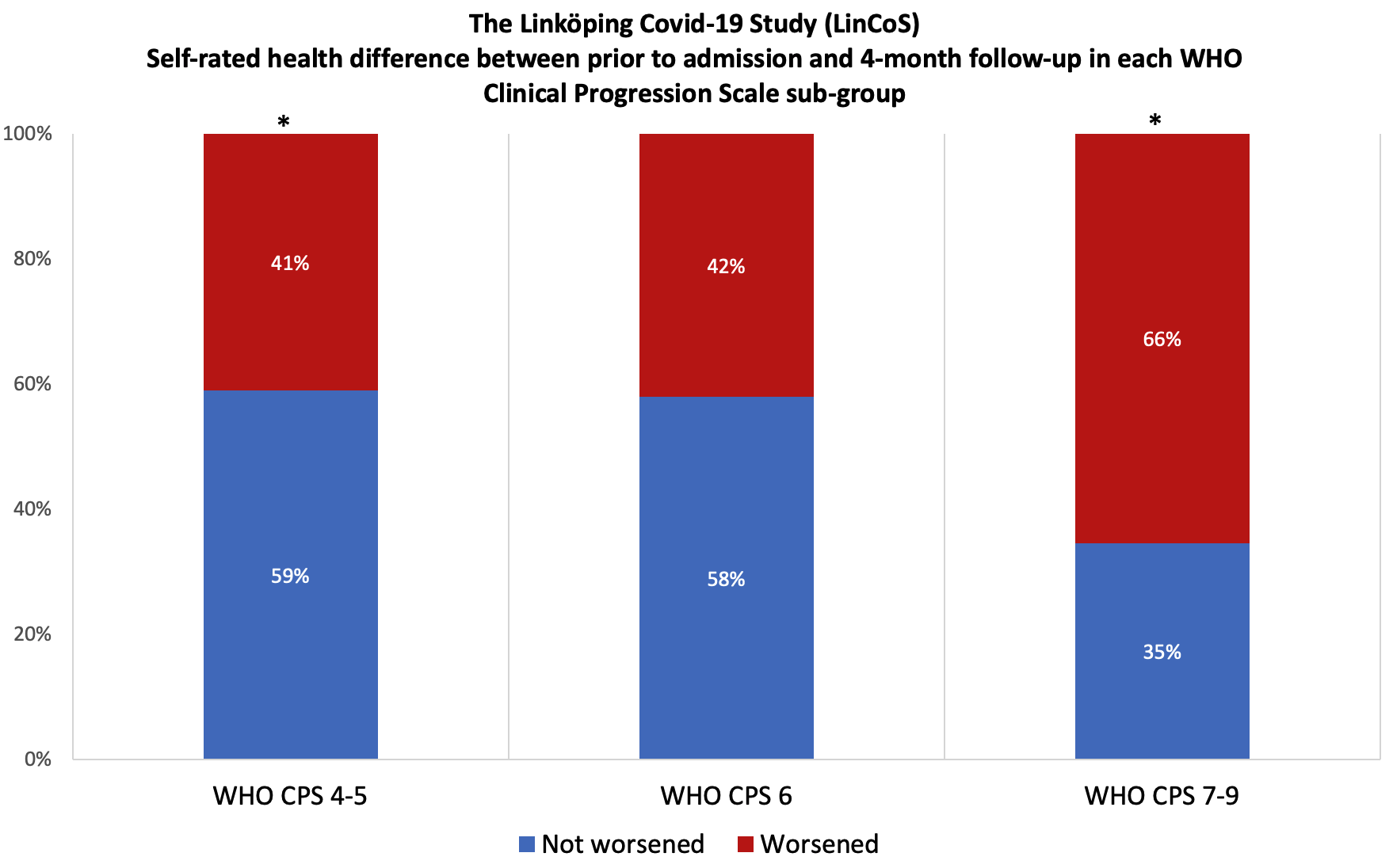

**Appendix II-Table 3.** Self-rated health difference between prior to admission and 4 months after discharge - Percentages of patients in each World Health Organization (WHO) Clinical Progression Scale (CPS) sub-group reporting. WHO CPS: grades 4-5 (hospitalised moderate disease, no oxygen or oxygen by mask or nasal prongs); grade 6 (hospitalised, severe disease, oxygen by NIV or high flow); grades 7-9 (hospitalised, severe disease, mechanical ventilation). *Indicates statistically significant difference between WHO CPS groups (p=0.003).

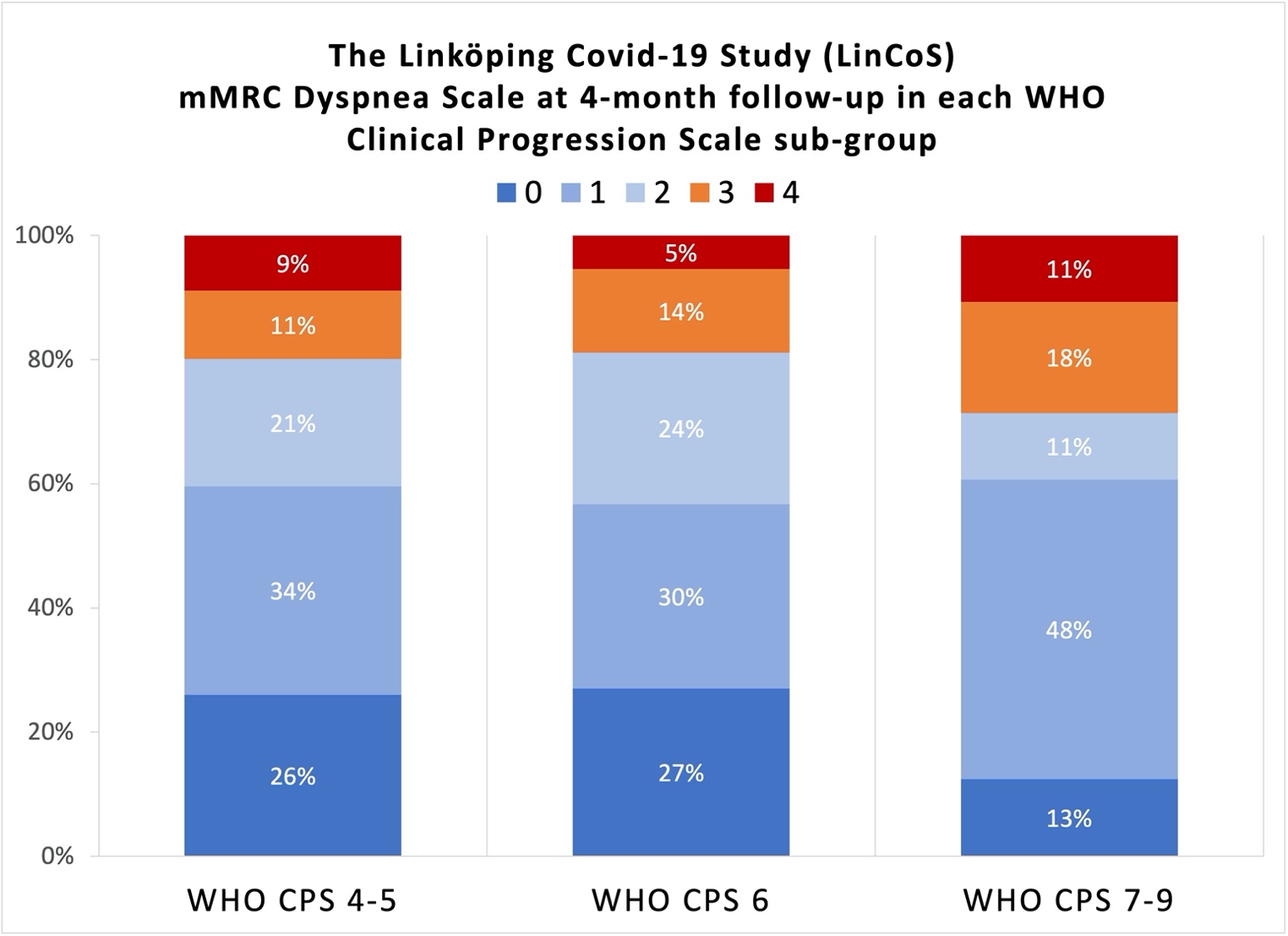

**Appendix II-Table 4.** Modified Medical Research Council (mMRC) Dyspnea Scale at 4 months after discharge - Percentages of patients in each World Health Organization (WHO) Clinical Progression Scale (CPS) sub-group reporting. WHO CPS: grades 4-5 (hospitalised moderate disease, no oxygen or oxygen by mask or nasal prongs); grade 6 (hospitalised, severe disease, oxygen by NIV or high flow); grades 7-9 (hospitalised, severe disease, mechanical ventilation). There were no statistically significant in-between group differences in regard to perceived breathlessness.
